# Antibody reactivity to SARS-CoV-2 is common in unexposed adults and infants under 6 months

**DOI:** 10.1101/2020.10.05.20206664

**Authors:** Abdelilah Majdoubi, Christina Michalski, Sarah E. O’Connell, Sarah Dada, Sandeep Narpala, Jean Gelinas, Disha Mehta, Claire Cheung, Manjula Basappa, Aaron C. Liu, Matthias Görges, Vilte E. Barakauskas, Jennifer Mehalko, Dominic Esposito, Inna Sekirov, Agatha N. Jassem, David M. Goldfarb, Daniel C. Douek, Adrian B. McDermott, Pascal M. Lavoie

## Abstract

**Background:** Pre-existing antibody reactivity against SARS-CoV-2 in unexposed people is a potentially important consideration for COVID-19 severity and vaccine responses. However, it has been difficult to quantify due to a lack of reliable defined background titers in unexposed individuals.

**Methods:** We measured IgG against multiple SARS-CoV-2 antigens, SARS-CoV and other circulating coronavirus spike proteins using a highly sensitive multiplex assay, and total SARS-CoV-2 spike-specific antibodies (IgG/M/A) using a commercial CLIA assay in 276 adults from the Vancouver area, Canada between May 17^th^ and June 19^th^ 2020. Reactivity threshold in unexposed individuals were defined comparing to pre-pandemic sera and to sera from infants under 6 months of age.

**Results:** The seroprevalence from a SARS-CoV-2 exposure, adjusted for false-positive and false-negative test results, was 0.60% in our adult cohort. High antibody reactivity to circulating endemic coronaviruses was observed in all adults and was ∼10-fold lower in infants <6 months. Consistent with a waning of maternal antibodies, reactivity in infants decreased more than 50-fold ∼8 months later. SARS-CoV-2 Spike, RBD, NTD or nucleocapsid antibody reactivity >100-fold above that of older infants was detected in the vast majority of unexposed adults and pre-pandemic sera. This antibody reactivity correlated with titers against circulating coronaviruses, but not with age, sex, or whether adults were healthcare workers.

**Conclusion:** A majority of unexposed adults have pre-existing antibody reactivity against SARS-CoV-2. The lack of similar antibody reactivity in infants where maternal antibodies have waned suggests that this cross-reactivity is acquired, likely from repeated exposures to circulating coronaviruses.

**Funding:** BC Children’s Hospital Foundation, NIH/NIAID

## Introduction

Coronavirus disease 2019 (COVID-19) was declared a global pandemic on March 11^th^, 2020 and has resulted in more than 43.6 million cases and 1.16 million deaths worldwide as of October 27, 2020. Almost all infected individuals seroconvert within 2-3 weeks, with the spike and nucleocapsid proteins eliciting the strongest responses (1, 2). While much attention has focused on SARS-CoV-2 responses in infected and convalescent individuals, the extent of SARS-CoV-2 antibody reactivity in unexposed individuals has been difficult to gauge due to insufficient assay sensitivity (3), or overlooked due to the lack of definable reactivity thresholds to distinguish reactive from unreactive unexposed individuals (4).

Four circulating coronaviruses pre-dating COVID-19 cause up to 30% of upper respiratory tract infections seasonally (5). The spike protein of circulating beta-coronaviruses HKU1 and OC43 exhibits ∼40% sequence similarity with SARS-CoV-2, whereas the circulating alpha-coronaviruses, NL63 and 229E, are less structurally similar (6). Since the emergence of SARS-CoV, MERS-CoV and SARS-CoV-2, interest in studying heterologous responses among human coronaviruses has increased due to their potential to explain why some individuals with COVID-19 have less severe outcomes (7, 8).

Across the world, estimates for SARS-CoV-2 seroprevalence range from 0.26-24.4%, with some reports suggesting higher rates among health care workers (HCW) (9, 10). In the province of British Columbia, Canada, there has been relatively little spread of SARS-CoV-2 during the first pandemic wave. British Columbia (BC, 5.017 million population) reported its first COVID-19 case on January 25. The first peak of the COVID-19 pandemic occurred between the third week of March and late April in BC (11). During this period, most economic sectors were locked down. With only 2,445 diagnosed COVID-19 cases (∼49/100,000 population) as of May 17, BC represented an ideal setting to compare the antibody reactivity against SARS-CoV-2 that can be generated from a direct viral exposure versus the pre-existing antibody reactivity against SARS-CoV-2 that may be detectable within the same unexposed population.

The main objective of this study was to determine the adult population-level antibody reactivity against SARS-CoV-2 and its relationship with seroreactivity to circulating coronaviruses. To confirm that SARS-CoV-2 antibody reactivity in unexposed adults was genuinely cross-reactive and not due to widespread unreported asymptomatic SARS-CoV-2 circulation, we similarly assayed sera collected prior to the emergence of SARS-CoV-2 and from infants before and after maternal antibodies have waned.

## Results

### Study population

In total, 276 adults were recruited. Their demographic characteristics and geographical area of residence are shown in **Supplemental Table 1** and **Supplemental Figure 1**. Majority (n = 196; 71%) of participants were HCW. Less than half had travelled outside of BC in 2020, including to the USA, Europe, Iran, the Caribbean, Australia, Mexico and Japan. Two participants reported a history of PCR-confirmed COVID-19.

**Figure 1.**
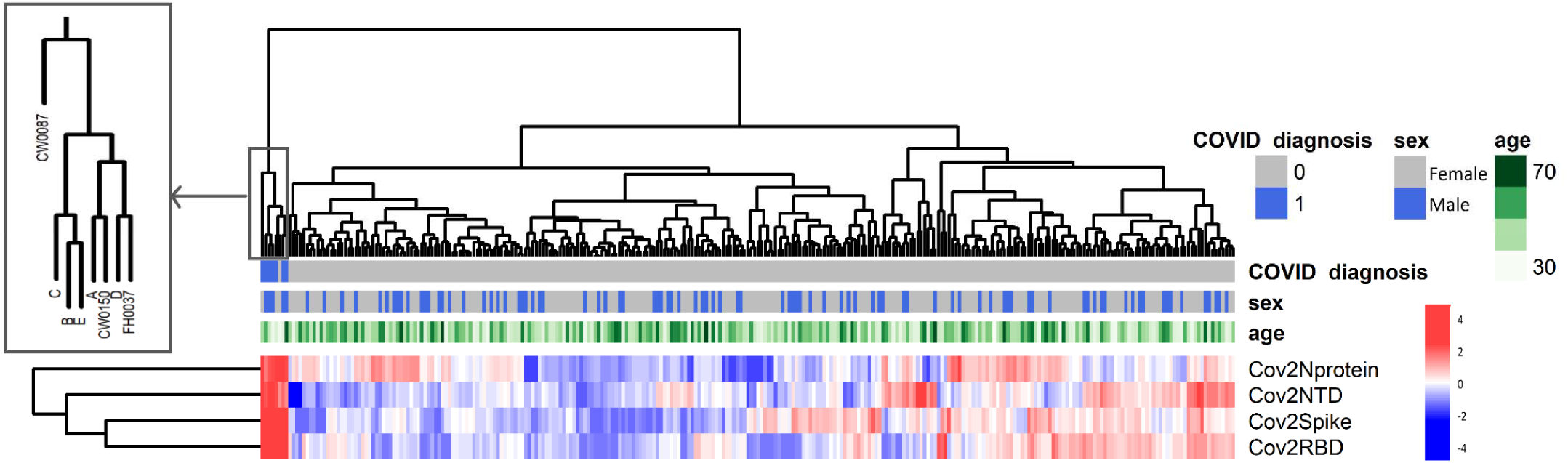
Hierarchical clustering analysis according to antibodies against SARS-CoV-2 antigens. This figure combines data from 276 study participants plus the five convalescent control sera. Colour scale represents antibody reactivity as a z-score.

### Prevalence of SARS-CoV-2 seroreactivity in adults

Using a highly sensitive multiplex assay, we quantified antibody titers against the spike proteins of circulating coronaviruses (OC43, HKU1, NL63, 229E), SARS-CoV and SARS-CoV-2 as well as against the N protein, RBD and NTD of SARS-CoV-2 from the 276 participants plus five control convalescent sera. Clustering analyses based on SARS-CoV-2 antibody profiles showed three individuals (CW087, CW0150, FH0037) that clustered very distinctly together with the five control convalescent sera, compared to the rest of the cohort (**Figure 1**). These three study participants included the two with a history of COVID-19 and one additional study participant: a 24-year-old asymptomatic non-HCW female who had been in contact with a COVID-19 case about 90 days prior to serology testing for this study.

Of the 276 participants, a subset of 222 that displayed above-the-mean antibody reactivity for any of the four SARS-CoV-2 antigens were tested with the highly specific CLIA assay (**Supplemental Figure 2**). The same 3 participants above, plus the 5 control sera, tested positive on this assay. Thus, we established that three out of 276 (1.1%) participants showed evidence of a prior SARS-CoV-2 exposure. After adjustment for bias using point estimates of specificity and sensitivity of the CLIA assay, the seroprevalence from a direct exposure to SARS-CoV-2 was established at 0.60% [95%CI 0% to 2.71%].

**Figure 2.**
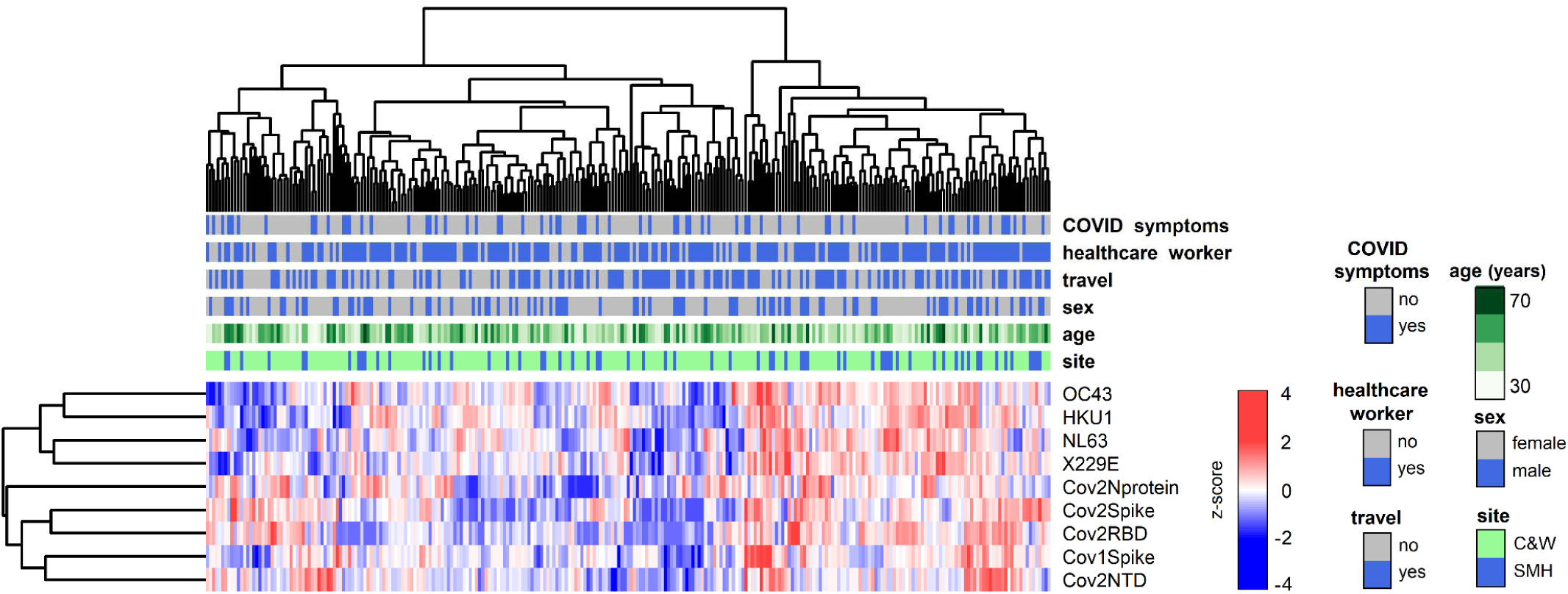
Hierarchical clustering based on antibody levels against 4 SARS-CoV-2, SARS-1 Spike antigens and human circulating coronaviruses from the 273 unexposed study participants by distribution of sociodemographic data. Colour scale represents antibody reactivity as a z-score.

The clinical characteristics of the SARS-CoV-2-seroreactive participants are shown in **Supplemental Table 2**. Two were HCW, one of whom was a pediatric HCW and the other was a non-pediatric HCW. Thus, there was no obvious difference in SARS-CoV-2 seroreactivity between HCW and non-HCW, or between pediatric and non-pediatric HCW (1.0% vs 1.0%; *P*=1.00 and 0.7% vs 1.6%; *P*=0.54, respectively), which may be indicative of similar rates of exposure to the virus in these groups in BC.

### Profile of SARS-CoV-2 antibody reactivity

The pattern of antibody reactivity against SARS-CoV-2 antigens differed markedly between convalescent sera and individuals who are seroreactive without any evidence of prior exposure to the virus. Indeed, individuals who had been previously infected with the virus showed high reactivity to *all* SARS-CoV-2 antigens tested (**Supplemental Figure 3A**), whereas unexposed individuals showed variable antibody reactivity against either Spike or RBD (**Supplemental Figure 3B, C**). For circulating coronaviruses, all individuals displayed high antibody reactivity (**Supplemental Figure 4**), which was evenly distributed by age, sex, HCW status, travel history and independent of participants’ prior impression of having experienced “COVID-19-like” symptoms (**Figure 2**). We detected significant correlations between antibody reactivities to SARS-CoV-2 and the circulating coronaviruses HKU1, NL63 and X229E (Spearman rho values ranging from 0.130 - 0.224 for 7 significant correlations with circulating coronaviruses out of 36 correlations tested) (FDR 5%; **Figure 3**).

**Figure 3.**
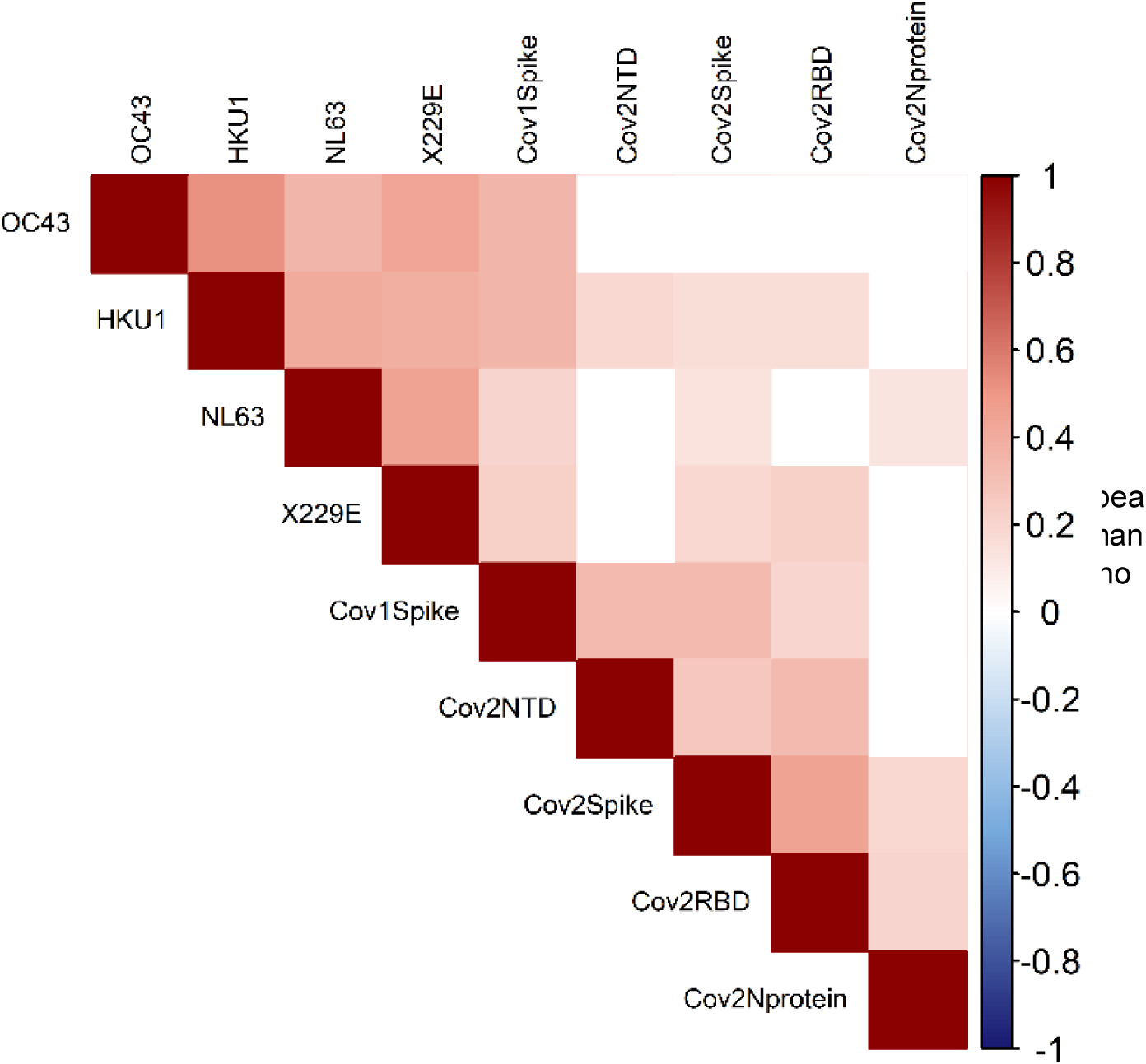
Correlations among SARS-CoV-2 and circulating coronaviruses antibody levels. Correlations among SARS-CoV-2 and circulating coronavirus antibody levels; Spearman rho values for significant correlations are shown on colour scale (FDR 5%; *P* values provided in Supplemental Table 3).

**Figure 4.**
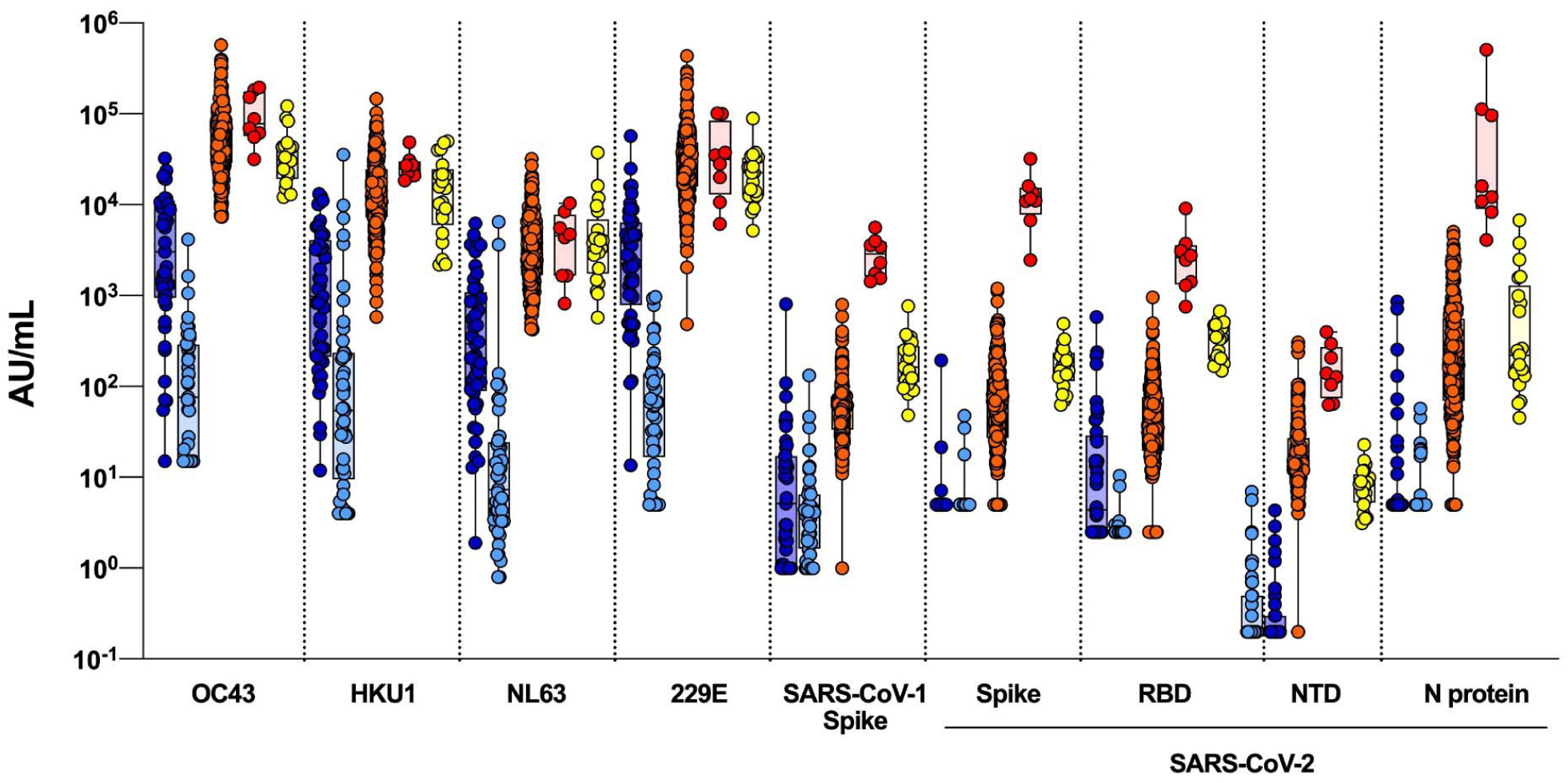
Comparison of serum antibody reactivity (Au/mL) between infants <6 months of age sampled before the COVID-19 pandemic (i.e. before Jan 1, 2020; darker blue) and again ∼8 months (lighter blue), SARS-CoV-2-unexposed adults (orange) and SARS-CoV-2-exposed adults (red) and pre-pandemic sera (yellow).

### SARS-CoV-2 antibody reactivity in unexposed individuals

Next, we aimed to define unequivocally the SARS-CoV-2 antibody reactivity thresholds in adults who have not been exposed to the virus. We reasoned that infants would be immunologically naïve with the exception of maternal antibodies that are expected to disappear gradually after birth. To this end, we performed the 10-plex assay on sera from 21 infants less than 6 months of age obtained before the pandemic and repeated in the same infants ∼8 months later, after BC’s lockdown period. Reactivity to circulating coronaviruses was almost uniformly detected in the first set of blood samples and this reactivity became substantially lower in the second sample sets (**Supplemental Figure 5**). Thus, these second infant samples allowed us to define a true threshold for background SARS-CoV-2-antibody reactivity (**Figure 4**). Most importantly, these data strikingly revealed that the vast majority of adults displayed some degree of antibody reactivity against either the S-2P native spike proteins, RBD, NTD or N proteins of SARS-CoV-2. SARS-CoV-2 reactivity in unexposed adults was up to ∼100-fold greater than infants after maternal antibodies had waned. Pre-pandemic sera also showed antibody reactivity comparable to unexposed study participants, therefore excluding that this reactivity was due to asymptomatic/undiagnosed exposures to the virus (**Figure 4**).

## Discussion

The goal of our study was to quantify the antibody reactivity against SARS-CoV-2 in the general population, particularly in uninfected individuals, and examine its relationship with reactivity against circulating endemic coronaviruses. We assessed three cohorts: adults sampled before the pandemic, a separate group of adults sampled during the pandemic, and infants sampled both before and during the pandemic. We estimated a 0.60% [95%CI 0% to 2.71%] seroprevalence from a direct SARS-CoV-2 exposure in the adult population. Reported SARS-CoV-2 seroprevalence after the first pandemic wave varied widely across the world for reasons including the timing of sampling; how countries managed to control spread of the virus; the target population of the study; and the assay used. Highest rates were reported in Sweden in May (7.3%) (12), Geneva in April/early May (10.8%) (13), and Madrid in late April/early May (>10%) (14); lowest rates were reported in BC (11). The findings from the current study are consistent with data from the BCCDC between May 15 and 27, 2020, reporting 0.55% seroprevalence among 885 residual sera obtained from an outpatient laboratory network in the Lower Mainland of BC; their study population represents a wider geographical catchment and did not specifically target HCW (11). The current study confirms that transmission of COVID-19 in BC after the first wave was low, even among HCW, contrasting with a high seroprevalence reported among HCW in other studies (10, 15, 16).

Antibody detection combining a commercial CLIA assay and the highly sensitive multiplex assay allowed us to distinguish antibody profiles between individuals who have been infected with SARS-CoV-2 from those who have not been exposed to the virus. This approach identified that a substantial proportion of “unexposed” individuals display antibody reactivity to SARS-CoV-2, but with a pattern quite distinct from that in convalescent individuals. The origin for this cross-reactivity is unclear. About 25% of participants in this study reported that they had experienced COVID-19-like symptoms. However, because the number of reported COVID-19 cases was low, despite an aggressive viral testing approach in BC (11), it is extremely unlikely that this antibody reactivity results from a direct exposure to SARS-CoV-2. Finding similar antibody reactivity in pre-pandemic sera also excludes this possibility.

The pre-existing SARS-CoV-2 antibody reactivity in unexposed individuals in the current study is consistent with T cell reactivity against SARS-CoV-2 detected in about 40% of unexposed individuals (17, 18). It is also consistent with another smaller study where 12 out of 95 pre-pandemic sera exhibited cross-reactive IgG antibody reactivity with conserved epitopes in SARS-CoV-2 proteins (S2 and N) (19). Previous seroprevalence studies have either used single-antigen assays, focused on SARS-CoV-2, or, in the latter case, examined cross-reactivity in selected sera (19). To the best of our knowledge, the current study is the first and largest to define background threshold to quantify the antibody reactivity against SARS-CoV-2 and circulating coronaviruses in SARS-CoV-2 unexposed adults, at the population level. Another strength of the study is that it reports antibody reactivity to SARS-CoV-2 and circulating coronaviruses in the same population, time period and assay, which is important to avoid sampling biases.

The correlations between the antibody reactivity to SARS-CoV-2 and circulating coronaviruses is consistent with 35-40% sequence similarity between the spike proteins of these two viruses (6). Correlations between HKU1, N63L and 229E, but not OC43 could suggest that unidentified factors modulate the extent of the cross-reactivity with different magnitude of cross-reactivity to circulating coronaviruses. Alternatively, it may simply due to chance, given our relatively small cohort. In contrast, it is not surprising that the antibody reactivity to SARS-CoV and SARS-CoV-2 is highly correlated given >75% sequence similarity between these two viruses (20). Importantly, reactivity detected on samples obtained from adults sampled before the emergence of COVID-19 and from young infants sampled also before the emergence of COVID-19, and again 8 months later confirms that we are detecting genuine heterologous reactivity rather than specific reactivity in exposed asymptomatic people. As a corollary it is possible that the antibody response following a COVID-19 episode is modulated by pre-existing immunity to circulating coronaviruses bolstered by an encounter with SARS-CoV-2.

This study has limitations. First, due to the lack of follow-up, findings from this study should not be used to predict whether the antibody reactivity to SARS-CoV-2 in unexposed individuals may confer any immune protection or harm. Second, infection with SARS-CoV-2 prior to 7 days may be underestimated by serology (21). However, this is unlikely to have had a significant effect on our estimates considering the low number of reported cases in BC. Third, the small sample size limited the power for correlation analyses and the precision of estimates of the overall prevalence from a direct SARS-CoV-2 exposure.

In conclusion, this study reveals that pre-existing antibody reactivity against SARS-CoV-2 antigens in sera from unexposed adults and in young infants is common, although it has been largely overlooked in previous serology studies. These findings warrant urgent studies of the impact such pre-existing seroreactivity may have on seroconversion following an exposure to SARS-CoV-2, likelihood of acquiring the infection, COVID-19 severity or vaccine responses.

## Methods

### Study design

Prospective cross-sectional study after the first pandemic wave in BC.

### Participants

Adults over 18 years of age from the greater Vancouver metropolitan area were included if they did not have *active* COVID-19, did not require self-isolation as per BC provincial public measures or had recovered from COVID-19 at least 14 days prior to the study visit and blood collection. Blood was drawn in gold-top serum separator tube with polymer gel (BD, cat# 367989); after at least 30 minutes of clotting at room temperature, the blood sample was then centrifuged at 1,400 G to obtain serum aliquots that were frozen at −80°C within four hours of collection. Infants were recruited from an ongoing study looking at respiratory syncytial virus antibodies, with enrollment/the first blood sample planned at the beginning and second blood sample at the end (after March 31^st^ 2020), of the winter viral season in BC. Residual de-identified sera from routine diagnostic testing performed in adults between January 25^th^ 2018 and November 8^th^ 2019 at the BCCDC Public Health Laboratory represent the pre-pandemic participants.

### Recruitment

Greater Vancouver is the main urban centre in BC and third largest metropolitan area in Canada with a population of 2.5 million. Study participants were invited by an emails sent to clinical departments of the BC Children’s & Women’s Hospitals (C&W, the largest pediatric referral centre in BC, located in Vancouver, and where no cases of COVID-19 were admitted during the pandemic first wave) and its affiliated BC Children’s Research Institute (BCCHR). The study was also advertised to hospitalists, anesthesiologists and critical care physicians at Surrey Memorial Hospital (located about 50 km from Vancouver). To minimize recruitment bias, all adults who responded to the invitation email and returned their signed consent form were enrolled sequentially and invited to give a blood sample, without triaging. Blood samples were collected between May 17 and June 19, 2020. Written informed consent was obtained from all participants. The study procedures were approved by the University of British Columbia (UBC) Children’s & Women’s Research Ethic Board (H20-01205; H18-01724).

### Study size

Since there was little population seroprevalence data available at the time and none in BC or Canada, no *a priori* sample size calculation was performed. The recruitment period was therefore defined by convenience over a three-week period of enrolment, in order to obtain baseline data.

### Multiplex antibody assay

A highly sensitive 10-plex assay (Meso Scale Diagnostics, Gaithersburg, USA) where each antigen is ‘spotted’ into a single well of a 96-well plate (22) was used to measure antibody profiles against four SARS-CoV-2 antigens: trimeric S-2P native spike protein, receptor-binding domain (RBD), N-terminal domain (NTD) (23) and nucleocapsid (N) protein; the trimeric SARS-CoV spike protein; and spike proteins from circulating beta- (HKU1, OC43) and alpha- (229E and NL63) coronaviruses, plus a negative control (bovine serum albumin, BSA), on all study participants plus 5 selected convalescent sera (A to E), included as positive controls (**Supplemental Figure 4**). Briefly, after blocking wells with 5% BSA, sera were added at 4 dilutions (2:25, 1:100, 1:800 and 1:10,000) and incubated with shaking for two hours. Sulfo-tag-labelled anti-IgG detection antibody were added and the electrochemiluminescence signal was read using the MSD Sector 600 instrument. Results are dilution-corrected interpolated values from a standard curve with assigned Arbitrary Units (AU)/mL. Assignment of AU/mL of serum was performed by Meso Scale Diagnostics and is designed such that values are comparable to an International Standard Serum (ISS), and that bridging to a WHO International Standard will be possible in the future.

### Commercial chemiluminescent (CLIA) antibody assay

Total antibody (IgA, IgG and IgM) against recombinant spike (S1) protein was determined using the VITROS 5600 analyser (Ortho-Clinical Diagnostics, Rochester, NY) according to manufacturer instructions. This is a Health Canada and FDA-licensed qualitative assay with reported performance and in-house validation indicating sensitivities >7 days post onset range between 96% and 100% and specificities from 99% to 100% (24, 25).

### Variables

The following information was collected from participants by questionnaire: age, sex, the first three digits of their postal code, health care workers status (and whether they worked at C&W or SMH), history of travel outside BC since January 1, 2020, and history of COVID-19 symptoms and testing. SARS-CoV-2-exposed cases were defined by a positive result on the commercial CLIA assay, validated for sensitivity by antibody profiling on the multiplex assay (detailed in Results).

### Statistical analyses

The seroprevalence for SARS-CoV-2 was calculated by dividing the number of SARS-CoV-2-exposed cases by the total number of participants, adjusted for bias due to false positive and false negative tests using the Greenland method (26). Differences in proportions were calculated using a Fisher exact test, with significance threshold at *P*<0.05. Hierarchical clustering of antibody levels (based on the multiplex assay), was performed on log-transformed, z-score normalized serology data, using the complete linkage agglomeration method and Euclidean distance measures. Spearman correlations between antibody levels and metavariables were adjusted for multiple testing using the Benjamini-Hochberg false-discovery rate method (FDR 5%). There were no missing data. Analyses were conducted in R version 4.0.2, R Studio version 3.6.2 and GraphPad Prism version 8.4.

## Data Availability

Non-identifying metadata will be made public (in process); participant-specific demographic data may be made available later, or to external investigators upon request, if can be agreed by participants and after obtaining permissions from relevant institutional ethics review boards...

## Author’s contributions

AM, CM and SD coordinated the study sample accrual and blood processing in Vancouver. JG and DM coordinated recruitment at SMH. CC collated the data and helped with data analysis. SEOC performed the multiplex assay, with help from SN and MB. AL performed the multiplex assay on the pre-pandemic adult sera. MG provided important input into the study design. ANJ and IS provided the BCCDC pre-pandemic sera. VEB and DMG supervised the commercial CLIA testing of samples. PML and ABM supervised the study in Vancouver and at the NAID/NIH, respectively. AM, CM, SD, DCD and PML wrote the manuscript first draft. All authors contributed to the study concept, design, data analysis and reviewing the manuscript and accept the article submission in its final form.

## Acknowledgements

We thank Lauren Muttucomaroe, Esther Alonso-Prieto and Lisa-Marie Candeias for help with recruitment; Linda Warner and Stuart Turvey for lending essential staff resources; Kim Schmidt and Kiara Gibbons for advertising the study; Mike Irvine for statistical consultations; and study participants who have generously donated their time and blood samples. AM is supported by a Mining for Miracles BCCHRI post-doctoral award. CC is supported by BCCHRI and UBC Faculty of Medicine Summer Student Research Program Studentships. AL is supported by a UBC Four Year Fellowship. PML holds a BCCHRI Award. ANJ acknowledges funding from the Michael Smith Foundation for Health Research.

---

**Supplemental Figure 1.**
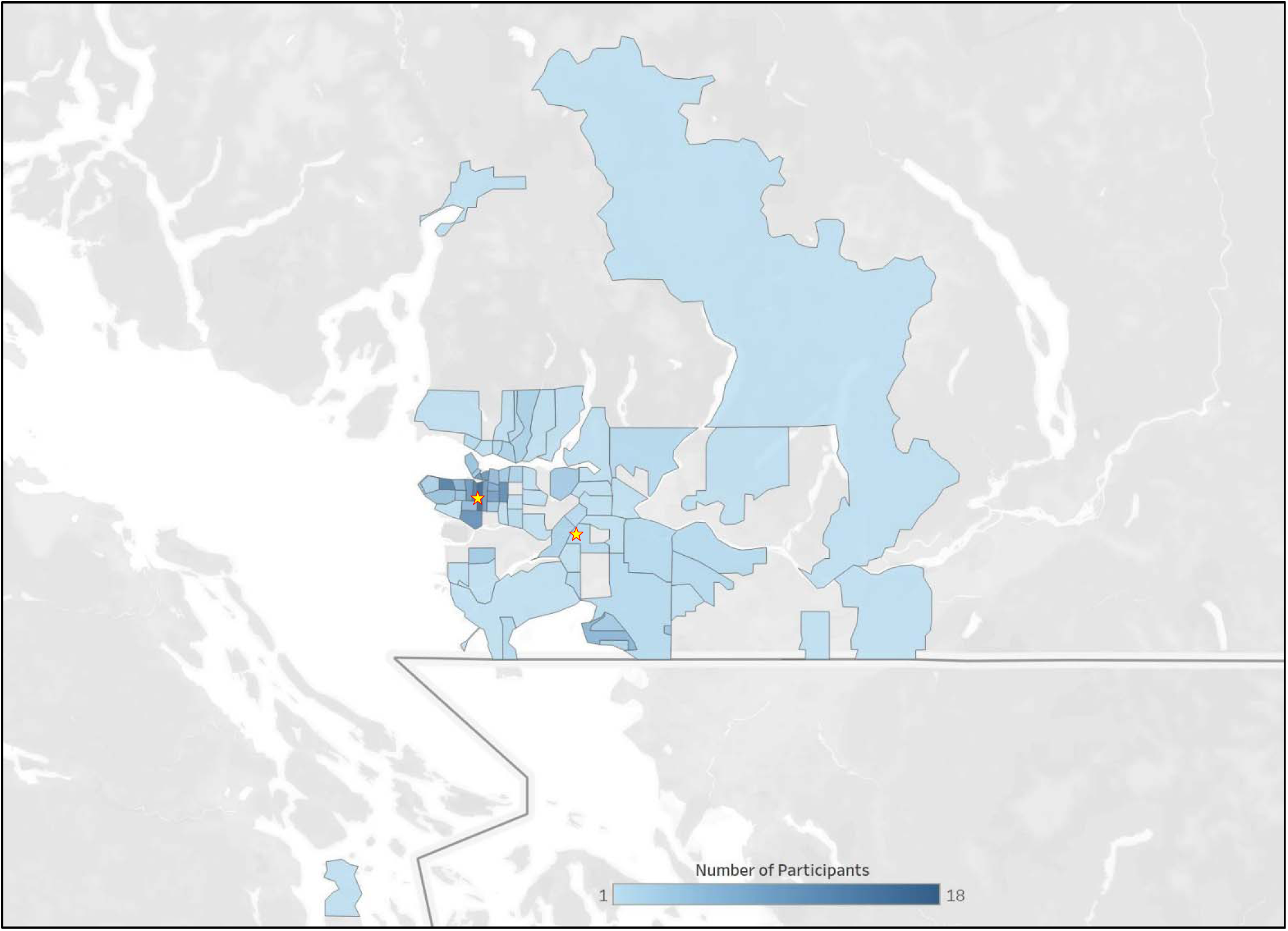
Geographical distribution. Data are from 276 study participants, based on postal code information. Stars indicate the location of the two recruitment sites (BC Children’s Hospital Research Institute on the left and Surrey Memorial Hospital on the right).

**Supplemental Figure 2.**
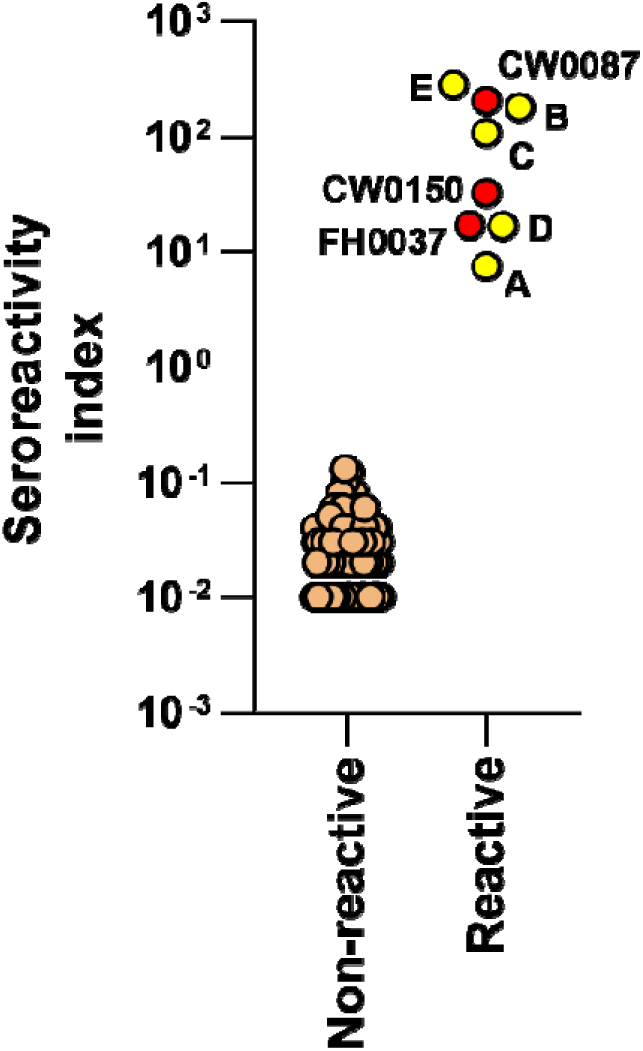
**Seroreactivity on the commercial CLIA assay** among 222 of 273 unexposed individuals who tested above the mean for at least one of the four SARS-CoV-2 antigens in the multiplex ECLIA assay, identifies three reactive sera (red) and 219 non-reactive sera (orange) in the study cohort. Data also shows seroreactivity indexes in the five convalescent control sera (yellow), for comparison.

**Supplemental Figure 3.**
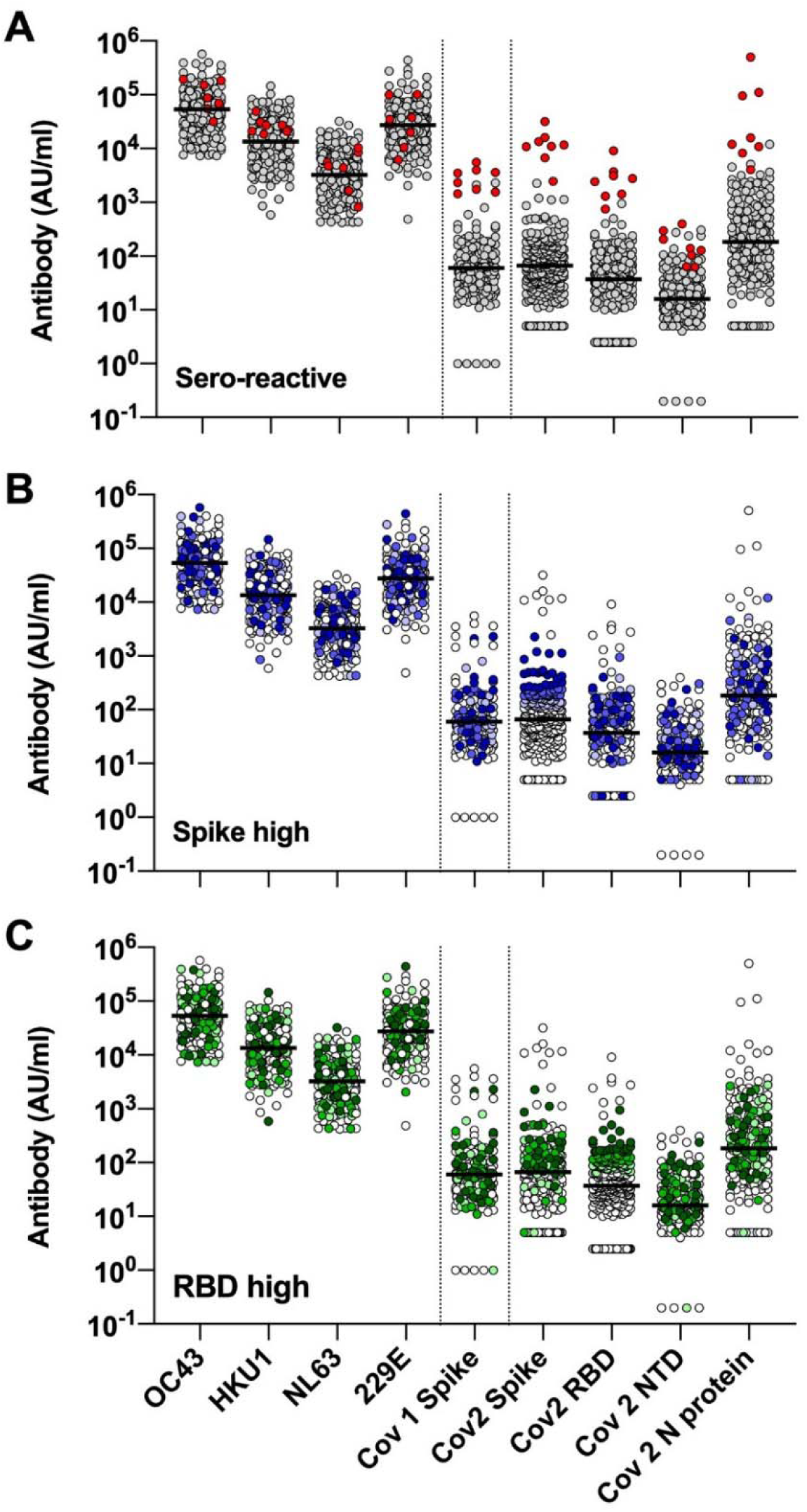
**Antibodies levels against circulating coronavirus and SARS antigens** in the combined group of 273 unexposed individuals, plus convalescent control sera, highlighting (**A**) SARS-CoV-2-exposed individuals (red), and (**B, C**) 273 unexposed participants with antibody reactivity against Spike or RBD at the top 90^th^ (darker tone), 80^th^ (mid-tone) or 70^th^ (lighter tone) centiles. Line = median. Null values are shown at the detection limit.

**Supplemental Figure 4.**
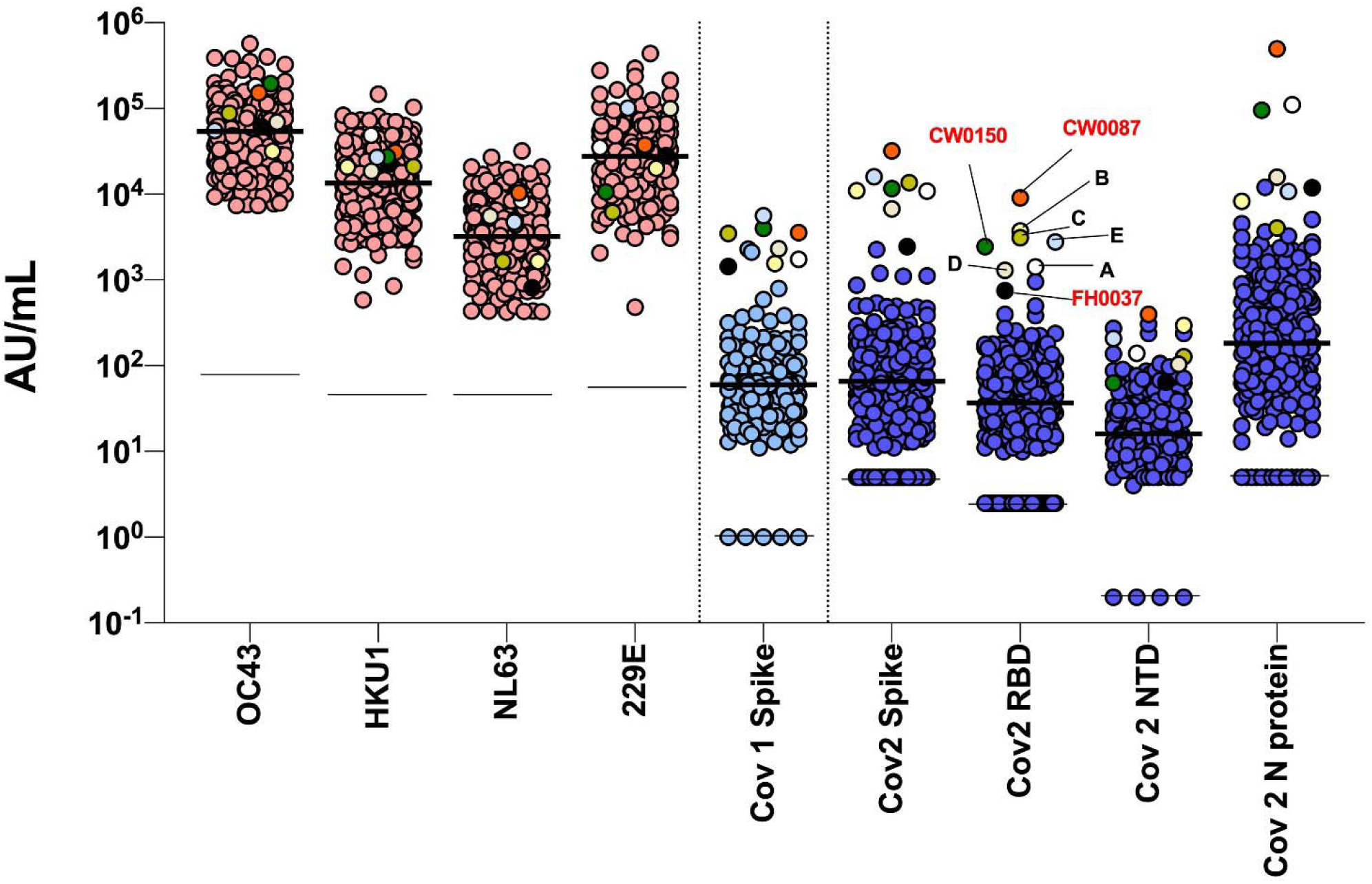
**Seroreactivity on the multiplex assay** among 276 study participants (including three exposed seroreactive participants), plus five convalescent control sera (A-E). The eight total reactive sera are multicolour-coded, whereas antibody levels are also grouped according to whether they represent circulating coronaviruses (pink), SARS-CoV (light blue) and SARS-CoV-2 antigens (dark blue). Thick black line = median. Thin black line = lower detection limit. Null values are shown at the detection limit.

**Supplemental Figure 5:**
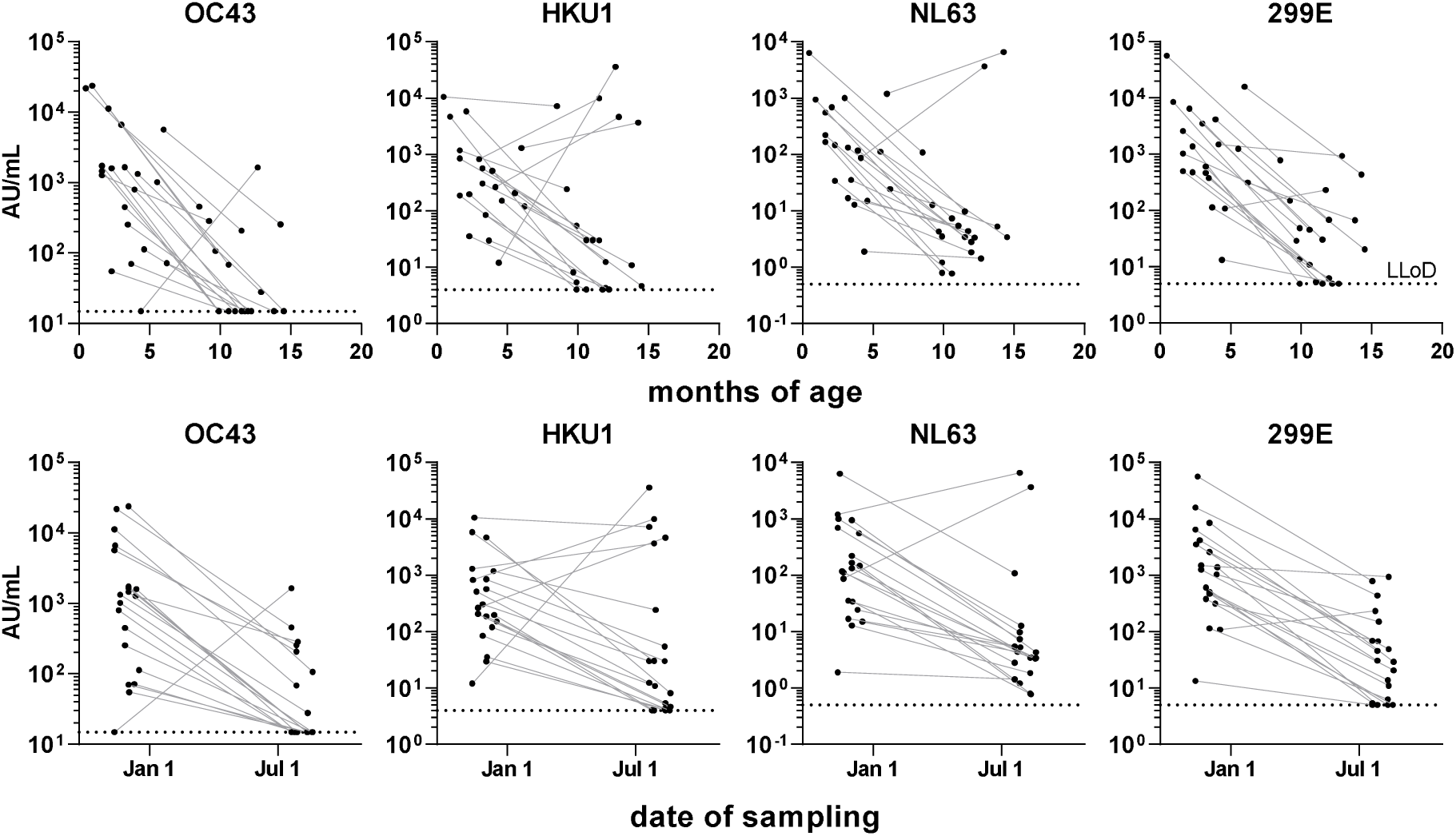
Paired antibody reactivity data for circulating coronaviruses in infants <6 months of age sampled before Jan 1, 2020 and ∼8 months later after the first pandemic wave and lockdown period in BC, by post-natal age (months) or date of sampling (2020); dotted line represents lower limit of detection (LLoD).

**Supplemental Table 1.**
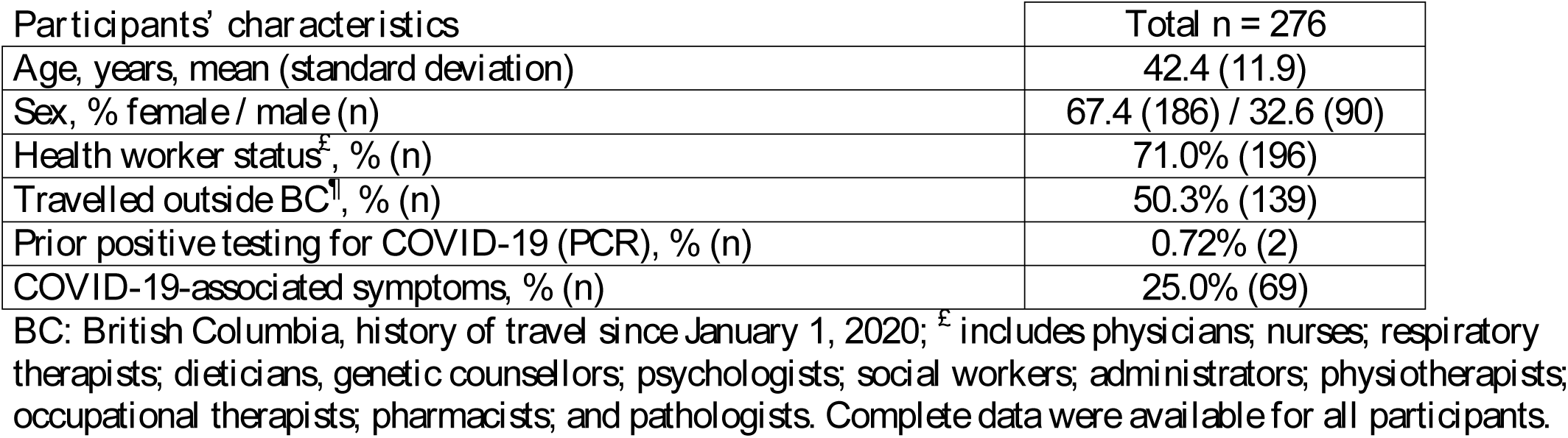
Demographical characteristics of study participants.

**Supplemental Table 2.** Clinical characteristics of three seroreactive study participants, plus five convalescent control sera.

| Participant ID# | Age | Sex | Travel outside BC | Convalescence period (days)* | Reported COVID-19-associated symptoms (by open-ended question) |
| --- | --- | --- | --- | --- | --- |
| CW0087<sup>£</sup> | 39 | Female | No | 57 | Nasal congestion, nasal dripping, laryngitis, intermittent dry cough, fatigue, chills, anosmia/ageusia with gradual return over 3 months |
| CW0150<sup>£</sup> | 24 | Female | No | unknown<sup>¶</sup> | Asymptomatic |
| FH0037<sup>£</sup> | 68 | Male | Yes | 72 | Shortness of breath on exertion, generalized aching, fever, mild cough |
| A<sup>€</sup> | 30 | Female | No | 75 | Cough, rhinorrhea, generalized body ache, extreme fatigue, head congestion, fever, anosmia/ageusia |
| B<sup>€</sup> | 35 | Male | Yes | 100 | Fatigue, severe headache/body aches, shortness of breath, drenching night sweats, mild diarrhoea, very mild dry cough, pain in both feet persisting |
| C<sup>€</sup> | 51 | Male | Yes | 96 | Protracted cough, fatigue, mild coryza and conjunctivitis, head congestion, and mild sore throat |
| D<sup>€</sup> | 28 | Male | No | 104 | Mild shortness of breath on exertion |
| E<sup>€</sup> | 25 | Male | No | 158 | Sore throat, headache, rhinorrhea, head congestion, muscle aches, severe fatigue, anosmia/ageusia (persisting >6 months), orthostatic syncope |

<sup>£</sup>Seroreactive study participants; <sup>€</sup>COVID-19 convalescent sera; \*time between COVID-19 diagnosis by PCR and serology testing.
<sup>¶</sup>Retrospectively (after participant was made aware of positive serology testing) we were able to identify that she had a contact with a COVID-19 case 89 days prior to serology testing.

**Supplemental Table 3.** Spearman rho and false discovery rate-adjusted p values (FDR 5%) for correlations for antibody levels between SARS-CoV-2 and circulating coronaviruses.

|  | OC43 | HKU1 | NL63 | X229E | Cov-1 Spike | Cov-2 NTD | Cov-2 Spike | Cov-2 RBD | Cov-2 N protein |
| --- | --- | --- | --- | --- | --- | --- | --- | --- | --- |
| OC43 | 0 | 3.60E-18 | 6.28E-09 | 6.04E-13 | 6.28E-09 | 0.062626 | 0.211963 | 0.364879 | 0.596884 |
|  | **1** | **0.512986** | **0.355693** | **0.432947** | **0.355877** | **0.120348** | **0.078814** | **0.056223** | **0.032149** |
| HKU1 |  | 0 | 2.28E-11 | 2.51E-10 | 6.28E-09 | 0.003351 | 0.014455 | 0.012611 | 0.083366 |
|  |  | **1** | **0.405455** | **0.385529** | **0.355032** | **0.188397** | **0.157294** | **0.160958** | **0.111909** |
| NL63 |  |  | 0 | 1.35E-13 | 0.001065 | 0.246839 | 0.026441 | 0.211963 | 0.045531 |
|  |  |  | **1** | **0.444881** | **0.209192** | **0.072401** | **0.143556** | **0.078796** | **0.130119** |
| X229E |  |  |  | 0 | 0.000261 | 0.108618 | 0.003569 | 0.000441 | 0.087371 |
|  |  |  |  | **1** | **0.233069** | **0.101703** | **0.185536** | **0.224499** | **0.109685** |
| Cov-1 Spike |  |  |  |  | 0 | 7.49E-08 | 1.25E-07 | 0.001866 | 0.06245 |
|  |  |  |  |  | **1** | **0.331146** | **0.324344** | **0.199227** | **0.121377** |
| Cov-2 NTD |  |  |  |  |  | 0 | 1.82E-05 | 1.00E-07 | 0.092581 |
|  |  |  |  |  |  | **1** | **0.268994** | **0.327365** | **0.107155** |
| Cov-2 Spike |  |  |  |  |  |  | 0 | 1.35E-13 | 0.003544 |
|  |  |  |  |  |  |  | **1** | **0.446363** | **0.186518** |
| Cov-2 RBD |  |  |  |  |  |  |  | 0 | 0.001065 |
|  |  |  |  |  |  |  |  | **1** | **0.209634** |
| Cov-2 N protein |  |  |  |  |  |  |  |  | 0 |
|  |  |  |  |  |  |  |  |  | **1** |

FDR-adjusted p values are indicated in **black** Spearman rho coefficients are indicated in **red**

## Notes

**Competing interests:** The authors declare no conflicts of interest

**Funding Statement:** This study was funded by the BC Children’s Hospital Foundation (to PML) and the Intramural Research Program of the Vaccine Research Centre (VRC) at the National Institute of Allergy and Infectious Diseases (NIAID), National Institutes of Health (NIH; to ABM and DCD). The funders did not play a role in the design, planning, execution, analysis or publication of the study.

### Competing Interest Statement

The authors have declared no competing interest.

### Clinical Trial

This study is not a clinical trial

### Funding Statement

This study was funded by the BC Children's Hospital Foundation (to PML) and the Intramural Research Program of the Vaccine Research Centre (VRC) at the National Institute of Allergy and Infectious Diseases (NIAID), National Institutes of Health (NIH; to ABM and DCD). The funders did not play a role in the design, planning, execution, analysis or publication of the study.

### Author Declarations

The study procedures were approved by the University of British Columbia (UBC) Children's & Women's Research Ethic Board (H20-01205; H18-01724).

### Summary of Updates

1st revision: Spearman correlations among serological outcomes and with metadata were corrected in the abstract (with adjustments in Figure 2). Corresponding rho/p values are now provided in Supplemental Tables. 2nd revision: Corrected Dr. Barakauskas' last name. 3rd revision: Corrected typos in Narpala and O'Connell's last names. Oct 19 and Oct 20 revisions: add two key authors and shorten abstract wording. Nov 5 revision: added new infant data and include major changes aligned with interpretation of these data;

